# Factors associated with care home resident quality of life: Demonstrating the value of a pilot Minimum Data Set using cross-sectional analysis from the DACHA study

**DOI:** 10.1101/2024.05.30.24308190

**Authors:** Stephen Allan, Stacey Rand, Ann-Marie Towers, Kaat De Corte, Freya Tracey, Elizabeth Crellin, Therese Lloyd, Rachael E Carroll, Sinead Palmer, Lucy Webster, Adam Gordon, Nick Smith, Gizdem Akdur, Anne Killett, Karen Spilsbury, Claire Goodman

## Abstract

**Background:** To maintain good standards of care, evaluations of policy interventions or potential improvements to care are required. A number of quality of life (QoL) measures could be used but there is little evidence for England as to which measures would be appropriate. Using data from a pilot Minimum Data Set (MDS) for care home residents from the Developing resources And minimum dataset for Care Homes’ Adoption (DACHA) study, we assessed the construct validity of QoL measures and analysed factors associated with QoL. This was to demonstrate the value of the pilot MDS data and to provide evidence for the inclusion of QoL measures in a future MDS.

**Methods:** Care home records for 679 residents aged over 65 from 34 care homes were available that had been linked to health records and official care home provider data. In addition to data on demographics, level of needs and impairment, several questions about the social care- and health-related QoL of participants were completed through proxy report (ASCOT proxy-resident, ICECAP-O, EQ5D-5D-5L Proxy 2). Construct validity was assessed through testing hypotheses developed from previous research and QoL measure constructs using discriminant analysis. Multilevel regression models were developed to understand how QoL was influenced by personal characteristics (e.g. sex, levels of functional and cognitive ability), care home level factors (type of home, level of quality) and resident use of health services (potentially avoidable emergency hospital admissions). Multiple imputation was used for missing data.

**Results:** All three measures were negatively associated with levels of cognitive impairment, whilst ICECAP-O and EQ-5D-5L Proxy 2 were negatively associated with low levels of functional ability. ASCOT Proxy-Resident was positively associated with aspects of quality and care effectiveness at both resident- and care home-level. All three QoL measures had acceptable construct validity and captured different aspects of QoL.

**Conclusion:** The study found acceptable construct validity for ASCOT-Proxy-Resident, ICECAP-O and EQ-5D-5L Proxy 2 in care homes as complementary measures based on different constructs. The study has demonstrated both the value of the DACHA study pilot MDS data and a rationale for the inclusion of these QoL measures in any future MDS.

## Background

Although global policy is progressively shifting towards provision of care in the community, many people still live in care homes. In England, around 315,000 people aged 65 and over live in care homes [1]. Limited budgets means that decisions need to be made, not only on where care is received, but also on how to maintain standards of care for people living in care homes, ensuring both quality of life [QoL] and good outcomes. Evaluations of policy or potential improvement in care are required, including economic evaluations.

An important outcome measure for care home residents is their QoL, which can be appropriately assessed using several measures. In terms of economic evaluation, it is ideal if these measures can be converted, along with knowledge of life expectancy, into quality-adjusted life years (QALYs), or an equivalent [2]. The most used measure of this kind is the EuroQol five-dimension questionnaire (EQ-5D), a measure of health-related quality of life [3]. However, given the aim of care is to support residents’ QoL, beyond health, it is important to also consider other measures of broader QoL for use in care home economic evaluations [4]. These include the Adult Social Care Outcomes Toolkit (ASCOT) and the ICEpop CAPability measure for older people (ICECAP-O). ASCOT is a suite of social care related QoL measures and is suitable for use in care homes [5-7]). ICECAP-O is a measure of QoL from a capability wellbeing perspective, assessing whether a person can do the things that are important to them in life [8-9]. ASCOT, which has been developed to be responsive to changes in social care delivery, and ICECAP-O, which looks at broader aspects of life in general for older people, may be preferable to EQ-5D for evaluations of care home interventions, as they could have greater sensitivity to QoL changes attributable to social care [10-11]. The value of using one QoL measure over another may depend on the focus of evaluation, e.g. health, social care, or life in general [12-13].

In England, care home resident QoL is positively associated with resident cognitive functioning, care home quality ratings and weakly with resident functional ability, when measured using ASCOT [7, 14-15]. The latter finding was expected given the instrument measures social care-related QoL, and, as such, a person’s (in)ability to achieve activities of daily living (ADLs) should be accounted for by the level of care they receive. ICECAP-O has been used in one previous study including residents in English care homes [16] with limited evidence on its validity [17]. Convergent and discriminant validity of the measure has been established for nursing home residents internationally, with both functional and cognitive ability associated with the measure [18-20]. There is limited evidence for EQ-5D-5L in English care homes, with international evidence associating the measure with functional ability [18,20] but not cognitive impairment [21-22], the latter indicating the difficulty of using a measure of health-related QoL in a care home setting.

For older people, in general, there is evidence on comparative differences between QoL instruments [10, 12, 23-24]. For care homes, there is emerging evidence of the feasibility and construct validity of the three QoL measures described above [25]. However, more evidence is required to consider the usefulness of QoL measures in understanding the needs, characteristics and health care utilisation of care home residents. In relation to health care utilisation, care home residents’ QoL is affected by hospitalisation, and particularly admissions deemed preventable [26-27]. As an indicator of unmet need, hospitalisation is an important outcome [28-29] and it is of interest to care home residents [30]. International studies have found that residents’ QoL is related to the quality of care received [31-32] and can predict risk of future hospitalisation [33-34]. However, research in English care homes is limited.

The Developing resources And minimum data set for Care Homes’ Adoption (DACHA) study assessed the feasibility of a Minimum Data Set (MDS) for care home residents [35]. This included developing a pilot MDS and demonstrating the value of the MDS data. MDS data included personal (e.g. sex, level of functional need and cognitive impairment) and care home (e.g. type and quality rating) characteristics. The data also included measures of health care utilisation, such as avoidable emergency hospital admission, and QoL. The aims of this study were to: i) assess the construct validity of three QoL instruments (ASCOT-Proxy-Resident, ICECAP-O and EQ-5D-5L Proxy 2) using discriminant analysis and ii) understand the factors associated with QoL. This was to both demonstrate the value of the pilot MDS data and to provide evidence for the inclusion of QoL measures in a future MDS.

## Methods

This study is reported using the Strengthening Reporting of Observational Studies in Epidemiology (STROBE) cross-sectional reporting guidelines [36].

### Study design and participants

Our study recruited 996 residents from 45 care homes (residential and nursing homes) in three areas of England that used digital care planning software from two providers. All permanent residents were eligible to be included in the study with the exception of those that were at the end-of-life, as judged by care home staff. Study design and participant recruitment are reported in detail elsewhere [35, 37].

The Pilot MDS (wave 1) contained data for 727 residents [35]. Forty-eight residents (6.6%) were excluded from analysis because either their record did not include a valid Care Quality Commission (CQC) care home ID to indicate where they lived, which was required for matching to care home level data (n=31) or they were under the age of 65 (n=17). Analysis proceeded using data for 679 residents, referred to herein as ‘the study sample’. The study sample was intended to be representative of the areas that participating care homes were located [37], but we could not confirm this with available data. Instead, we assessed the national representativeness of the residents in the study sample to the 2021 care home resident population aged over 65 [1].

### Dependent variables

In addition to existing digital care records on personal demographics and needs, several QoL measures were selected for inclusion in the pilot MDS [38]. In particular, data were collected through staff proxy response for ASCOT Proxy-Resident and ASCOT Proxy-Proxy (social care-related QoL), ICECAP-O (capability wellbeing), QUALIDEM (dementia-specific QoL), and EQ-5D-5L Proxy 2 (health-related QoL). For this analysis, three QoL measures, ASCOT Proxy-Resident, ICECAP-O and EQ-5D-5L Proxy 2, were included as they were found to be feasible to collect, valid and internally consistent through evaluation of their psychometric properties [25, 39].

#### Ascot-Proxy-Resident

ASCOT-Proxy is a questionnaire collecting data for two separate measures of social care-related QoL (SCRQoL) [40]. It covers eight social care domains: personal comfort and cleanliness, personal safety, food and drink, activities/occupation, control over daily life, social participation, home cleanliness and comfort, and dignity, with four levels (ideal state, no needs, some needs, and high needs). Proxy respondents (in this study, care home staff) are asked to rate ASCOT-Proxy items from both the proxy-resident (i.e. what the proxy thinks the resident thinks) and proxy-proxy (i.e. what the proxy thinks about the resident’s QoL) perspective. Two measures of proxy-report social care-related QoL are generated: ASCOT-Proxy-Resident and ASCOT-Proxy-Proxy. ASCOT-Proxy-Resident was included in this analysis because it has been found to be a valid QoL instrument, with the same structure as the original ASCOT-SCT4, for data collected with proxy respondents for care home residents [39]. An index score (-.171 to 1) was generated using preference weights for ASCOT-SCT4, with 0 being equivalent to ‘being dead’ and 1 representing the ideal social care-related QoL state [5].

#### ICECAP-O

ICECAP-O is a measure of capability wellbeing [9, 41]. ICECAP-O has five items: attachment, security, role, enjoyment, and control, with four levels of response that represent capability (none, a little, a lot, and all). In this study, ICECAP-O was collected using proxy report and the score (0 to 1) was calculated using UK index values [9], ranging from 0 (no capability) to 1 (full capability).

#### EQ-5D-5L Proxy 2

The EQ-5D-5L measures individuals’ level of functioning in five domains: pain, mobility, usual activities, anxiety/depression, and self-care, with five levels (no problems, slight problems, moderate problems, severe problems, and extreme problems) [42-43]. In this study, we used the EQ-5D-5L Proxy 2 version, which asks proxy respondents (care staff) to rate QoL from the proxy-resident perspective. The EQ-5D-5L score (-.594 to 1) was calculated using the mapping function to convert to EQ-5D-3L and applying its UK index values since the UK value set for EQ-5D-5L is still being developed [44-45]. A score of 1 represents full health and 0 is an equivalent state to death.

### Independent variables

The pilot MDS contained data on a number of factors likely to be associated with QoL which were included in the analysis. The following measures of personal function were included: functional independence (Barthel Index), cognitive impairment (MDS Cognitive Performance Scale, MDSCPS) and delirium (Informant Assessment of Geriatric Delirium Scale, IAGeD) [46-48]. Length of stay was calculated using date of admission from care home records. Sex, age, and ethnicity were available from health records. Given the very small number of residents that were not in the White high-level ethnic grouping (n=8), we excluded ethnicity from the regression analysis. Comorbidities was measured if a resident had two or more Elixhauser comorbidities during hospital admissions in the previous three years [49-50], and potentially avoidable emergency hospital admissions in the previous 12 months was included in the study as an indicator of unmet needs. A potentially avoidable admission was where the primary diagnosis was a condition considered manageable, treatable or preventable in community settings, or that may be caused by poor care or neglect [51]. Finally, we included the following factors at a care home level: type of home, number of beds, occupancy rate, self-funding rate (i.e. percentage of residents funding their own stays) and most recent CQC quality rating.

Resident-level independent variables that were continuous or counts were recoded into categories based on previous research and measure constructs [15,19,48].

### Hypotheses

To establish construct validity by hypothesis testing, we used discriminant analysis and tested for differences in QoL score between residents dependent on characteristics. Table 4 presents the hypotheses, which were based on previous research or *a priori* informed by the measurement constructs. Sufficient evidence of construct validity was considered using a criterion of >=75% of hypotheses accepted [52].

### Statistical methods

#### Missing data strategy

There was missing data in the MDS. We confirmed that the data were not missing completely at random (MCAR) using logistic regression of binary missing data indicators for each QoL measure. Therefore, we assumed the data were missing at random (MAR) and used multiple imputation (MI) to address the missing data given a complete case analysis could provide inefficient and biased estimates [53]. The QoL models were multi-level (see below). As such, we imputed the data using the chained imputation method with predictive mean matching (QoL measures, age, length of stay, Barthel index, IAGeD, occupancy rate, self-funding rate, staff-resident ratio), Poisson (number of comorbidities) and ordered logistic (MDSCPS) models at two levels, care home and resident [54-55]. In the first imputation step, we imputed missing care home level data (ten imputations), using care home means of known resident characteristics [54,56]. We then included each care home level imputation individually in the imputation of data at the resident level (ten imputations). This generated a resident level dataset with 100 imputations. We conservatively chose the number of imputations to provide adequate levels of reproducibility (i.e., the same results would be found if the multiple imputation was repeated). To confirm reproducibility of the findings, we assessed the random errors generated by the MI process in the estimations of QoL [57].

#### Regression model

To allow for factors that could influence the variation in QoL at resident and care home level, we estimated a ‘within-between’ multi-level model of each QoL measure, with care home residents clustered by care home [58] and categorical variables included in models using dummy codes [59]. The within-between multi-level model separates out level 1 (i.e., resident) associations into within and between associations, and, at the same time, allows for appropriate estimation of other care home characteristics and controls for omitted variables at the care home level (e.g. location).

Given MI, the appropriateness of the multi-level structure was assessed pragmatically with Likelihood-ratio tests of the null hypothesis of no variance between care homes for each imputation. Standard errors were clustered by care home.

We carried out the analysis using Stata 18 SE version and set a statistical significance using two-sided tests of 0.05.

## Results

### Descriptive statistics

Representativeness of the study sample to the overall care home resident population of England is presented in Table 1. The study sample was similar to the resident population by sex and type of care home but overrepresented the very old and the White high-level ethnic grouping. Table 2 presents descriptive data of the residents in the study sample, including levels of missing data. Construct validity by hypothesis testing that considered correlations between the three measures is reported elsewhere [25]. The complete data showed that the average resident had an ASCOT-Proxy-Resident, ICECAP-O and EQ-5D score of 0.831, 0.738 and 0.342, respectively. There was a high level of need amongst the residents, as 70% of residents had low functional independence, whilst cognitive impairment as assessed by MDSCPS ranged from no impairment (19.0%) to severe or very severe impairment (25.4%). A number of residents (16.3%) had a potentially avoidable emergency hospital admission in the previous year. The residents lived in 34 care homes, 19 of which were nursing homes. The majority of the homes were rated as ‘Good’ (70.6%), with 17.6% rated ‘Requires improvement’ and 11.8% ‘Outstanding’. No homes were rated as ‘Inadequate’. The average care home in the sample had 48 beds, an occupancy rate of 87.8%, and almost half of all residents self-funding, i.e. paying for their own care.

**Table 1:** Comparison of study sample to English care home resident population.

|  | Study sample | Care home resident population | Study sample v population (t-test) |
| --- | --- | --- | --- |
| Age 85 and over (%) | 60.0 | 56.4 | 2.04* |
| Non-white (%) | 1.2 | 2.5 | -2.99** |
| Male (%) | 28.9 | 30.3 | -0.82 |
| Nursing home (%) | 51.8 | 49.4 | 1.27 |

Source: Care home resident population data taken from 2021 census data (ONS, 2023). \*p < .05, \*\* p < .01.

**Table 2:**
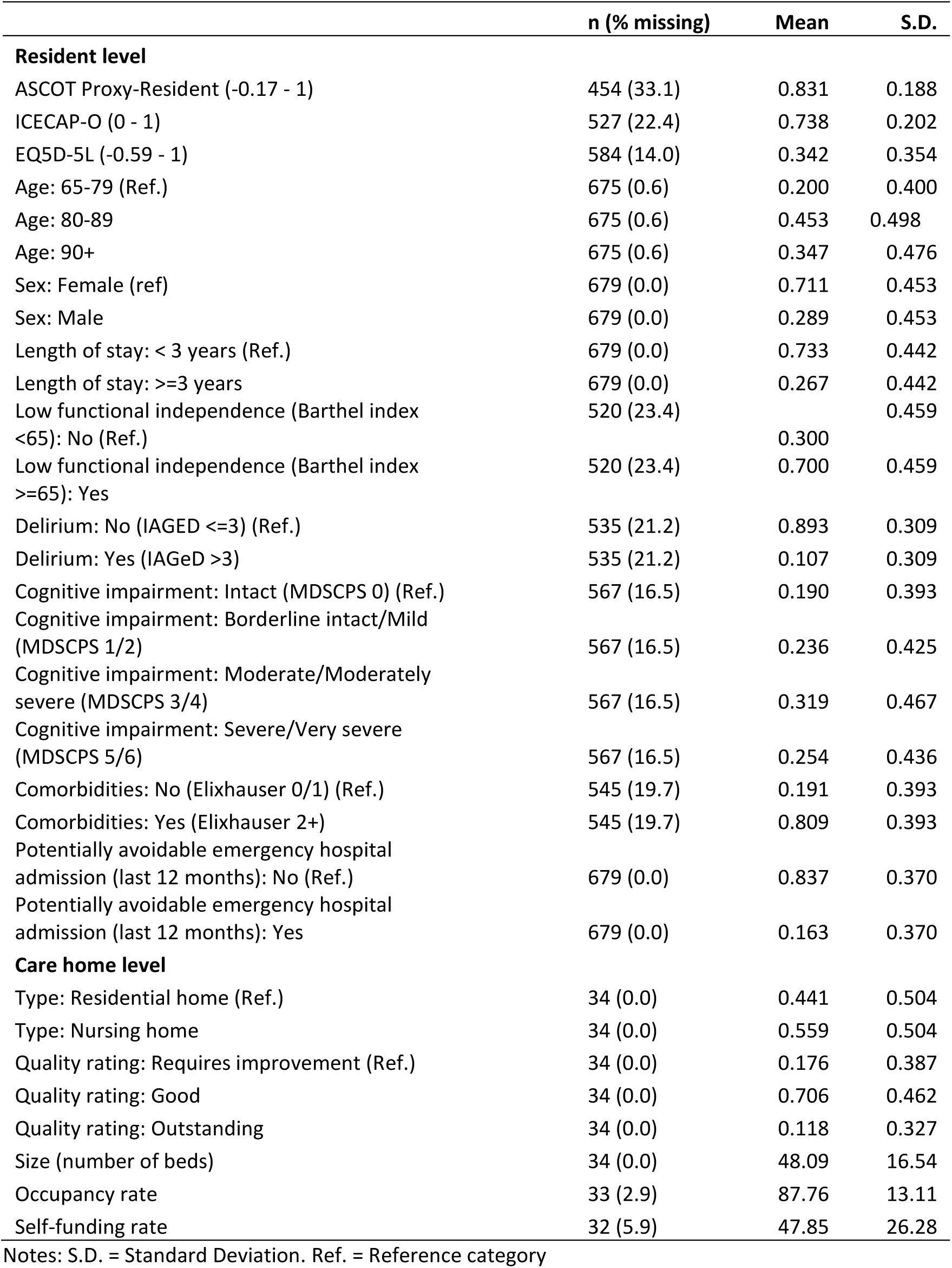
Descriptive statistics of the study sample.

|  | n (% missing) | Mean | S.D. |
| --- | --- | --- | --- |
| **Resident level** |  |  |  |
| ASCOT Proxy-Resident (-0.17 - 1) | 454 (33.1) | 0.831 | 0.188 |
| ICECAP-O (0 - 1) | 527 (22.4) | 0.738 | 0.202 |
| EQ5D-5L (-0.59 - 1) | 584 (14.0) | 0.342 | 0.354 |
| Age: 65-79 (Ref.) | 675 (0.6) | 0.200 | 0.400 |
| Age: 80-89 | 675 (0.6) | 0.453 | 0.498 |
| Age: 90+ | 675 (0.6) | 0.347 | 0.476 |
| Sex: Female (ref) | 679 (0.0) | 0.711 | 0.453 |
| Sex: Male | 679 (0.0) | 0.289 | 0.453 |
| Length of stay: < 3 years (Ref.) | 679 (0.0) | 0.733 | 0.442 |
| Length of stay: ≥3 years | 679 (0.0) | 0.267 | 0.442 |
| Low functional independence (Barthel index <65): No (Ref.) | 520 (23.4) | 0.300 | 0.459 |
| Low functional independence (Barthel index ≥65): Yes | 520 (23.4) | 0.700 | 0.459 |
| Delirium: No (IAGED ≤3) (Ref.) | 535 (21.2) | 0.893 | 0.309 |
| Delirium: Yes (IAGeD >3) | 535 (21.2) | 0.107 | 0.309 |
| Cognitive impairment: Intact (MDSCPS 0) (Ref.) | 567 (16.5) | 0.190 | 0.393 |
| Cognitive impairment: Borderline intact/Mild (MDSCPS 1/2) | 567 (16.5) | 0.236 | 0.425 |
| Cognitive impairment: Moderate/Moderately severe (MDSCPS 3/4) | 567 (16.5) | 0.319 | 0.467 |
| Cognitive impairment: Severe/Very severe (MDSCPS 5/6) | 567 (16.5) | 0.254 | 0.436 |
| Comorbidities: No (Elixhauser 0/1) (Ref.) | 545 (19.7) | 0.191 | 0.393 |
| Comorbidities: Yes (Elixhauser 2+) | 545 (19.7) | 0.809 | 0.393 |
| Potentially avoidable emergency hospital admission (last 12 months): No (Ref.) | 679 (0.0) | 0.837 | 0.370 |
| Potentially avoidable emergency hospital admission (last 12 months): Yes | 679 (0.0) | 0.163 | 0.370 |
| **Care home level** |  |  |  |
| Type: Residential home (Ref.) | 34 (0.0) | 0.441 | 0.504 |
| Type: Nursing home | 34 (0.0) | 0.559 | 0.504 |
| Quality rating: Requires improvement (Ref.) | 34 (0.0) | 0.176 | 0.387 |
| Quality rating: Good | 34 (0.0) | 0.706 | 0.462 |
| Quality rating: Outstanding | 34 (0.0) | 0.118 | 0.327 |
| Size (number of beds) | 34 (0.0) | 48.09 | 16.54 |
| Occupancy rate | 33 (2.9) | 87.76 | 13.11 |
| Self-funding rate | 32 (5.9) | 47.85 | 26.28 |

Notes: S.D. = Standard Deviation. Ref. = Reference category

### Discriminant analysis

Table 3 presents the results of the multi-level regressions of QoL, as measured by ASCOT-Proxy-Resident, ICECAP-O and EQ-5D-5L. Likelihood-ratio tests for all three QoL measures found there was significant evidence of variation between care homes (ρ<0.01 for all 100 imputations), confirming the multi-level modelling strategy as appropriate. We found no evidence that the random errors generated by MI significantly affected the findings for any of the QoL regression models, confirming reproducibility if the MI process was repeated.

**Table 3:** Multi-level regression models predicting QoL (n=679)

| Parameters | Model 1: Ascot-Proxy-Resident |  | Model 2: ICECAP-O |  | Model 3: EQ-5D-5L Proxy 2 |  |
| --- | --- | --- | --- | --- | --- | --- |
|  | Coef. | S.E. | Coef. | S.E. | Coef. | S.E. |
| **Resident variables** |  |  |  |  |  |  |
| Sex: Male | 0.011 | 0.017 | -0.030 | 0.017 | 0.049 | 0.023 |
| Age: 80-89 | 0.026 | 0.018 | 0.018 | 0.019 | -0.007 | 0.026 |
| Age: 90+ | 0.040* | 0.020 | 0.016 | 0.019 | -0.029 | 0.026 |
| Length of stay: ≥3 years | -0.023 | 0.018 | -0.027 | 0.018 | -0.107*** | 0.021 |
| Low functional independence: Yes | -0.021 | 0.016 | -0.064*** | 0.015 | -0.342*** | 0.042 |
| Delirium: Yes | -0.067* | 0.031 | -0.057* | 0.026 | -0.007 | 0.033 |
| Cognitive impairment: Borderline intact/Mild | -0.037* | 0.017 | -0.028 | 0.016 | -0.020 | 0.025 |
| Cognitive impairment: Moderate/Moderate Severe | -0.078** | 0.023 | -0.075*** | 0.021 | -0.038 | 0.030 |
| Cognitive impairment: Severe/Very severe | -0.081** | 0.025 | -0.172*** | 0.023 | -0.177*** | 0.037 |
| Comorbidities: Yes | 0.008 | 0.019 | -0.008 | 0.020 | -0.034 | 0.028 |
| Potentially avoidable emergency hospital Admission: Yes | -0.044* | 0.020 | -0.001 | 0.019 | -0.043 | 0.029 |
| **Care home variables** |  |  |  |  |  |  |
| Sex: Male (Mean) | 0.160 | 0.159 | -0.190 | 0.246 | 0.159 | 0.248 |
| Age: 80-89 (Mean) | -0.091 | 0.160 | -0.122 | 0.226 | 0.648** | 0.206 |
| Age: 90+ (Mean) | 0.148 | 0.142 | 0.105 | 0.174 | -0.097 | 0.183 |
| Length of stay: ≥3 years (Mean) | -0.011 | 0.121 | -0.062 | 0.182 | 0.334 | 0.179 |
| Low functional independence: Yes (Mean) | -0.091 | 0.067 | -0.132 | 0.068 | -0.527*** | 0.135 |
| Delirium: Yes (Mean) | 0.160 | 0.146 | -0.049 | 0.167 | -0.221 | 0.173 |
| Cognitive impairment: Borderline intact/Mild (Mean) | 0.098 | 0.161 | -0.006 | 0.155 | 0.326 | 0.195 |
| Cognitive impairment: Moderate/Moderate Severe (Mean) | -0.056 | 0.096 | 0.028 | 0.120 | 0.343** | 0.125 |
| Cognitive impairment: Severe/Very severe (Mean) | -0.250 | 0.133 | -0.212 | 0.138 | -0.188 | 0.145 |
| Comorbidities: Yes (Mean) | -0.023 | 0.104 | 0.049 | 0.131 | -0.221 | 0.170 |
| Potentially avoidable emergency hospital Admission: Yes (Mean) | -0.098 | 0.196 | 0.171 | 0.171 | 0.064 | 0.196 |
| Type: Nursing home | -0.090* | 0.035 | -0.126* | 0.040 | -0.078 | 0.049 |
| Quality rating: Good | -0.023 | 0.050 | -0.027 | 0.068 | 0.022 | 0.077 |
| Quality rating: Outstanding | 0.113* | 0.056 | 0.134 | 0.085 | 0.036 | 0.089 |
| Size (number of beds) | 0.001 | 0.001 | 0.0017 | 0.001 | 0.0003 | 0.002 |
| Occupancy rate | -0.0026* | 0.001 | -0.0002 | 0.001 | 0.0001 | 0.001 |
| Self-funding rate | -0.001 | 0.001 | -0.001 | 0.001 | 0.0028* | 0.0014 |
| Number of imputations | 100 |  | 100 |  | 100 |  |
| Relative variance inflation (RVI) | 0.614 |  | 0.508 |  | 0.597 |  |
| Fraction of missing information (FMI) | 0.497 |  | 0.356 |  | 0.685 |  |

Note: \*p < .05, \*\*p < .01, \*\*\*p < .001. Reference categories are Sex: female; Age: 65-79; Length of stay: <3 years; Low functional independence: No; Delirium: No; Cognitive impairment: Intact; Emergency admission: No; Comorbidities: No; Type: Residential home; Quality rating: Requires improvement.

ASCOT-Proxy-Resident was significantly negatively influenced by level of cognitive impairment and previous potentially avoidable emergency hospital admission and positively influenced by living in a care home with an ‘Outstanding’ CQC quality rating. There was no significant influence of functional dependence on ASCOT-Proxy-Resident. In contrast, ICECAP-O was significantly negatively influenced by functional dependence, and also by level of cognitive impairment. Whereas ASCOT-Proxy-Resident could discern a difference in score at lower levels of impairment, ICECAP-O had differences in score between higher levels of cognitive impairment relative to having no impairment. In particular, within a care home, the average resident with moderate/moderately severe and severe/very severe impairment had a 0.075 and 0.172 lower ICECAP-O score than those with no impairment, respectively.

EQ-5D-5L was negatively influenced by functional dependence and the highest level of cognitive impairment, but not by previous potentially avoidable emergency hospital admission. The size of influence of low functional independence, for the average resident within a care home, was greatest on EQ-5D-5L (0.342 lower score, equivalent to 100% of average score) compared to both ICECAP-O (0.064, 8.9% of average score) and ASCOT (not significantly different from zero).

ASCOT-Proxy-Resident score was lower for those aged 90 years and over and residents with delirium. It was also lower for residents living in nursing homes and homes with higher occupancy levels. ICECAP-O score was significantly lower for residents with delirium and those in nursing homes, and EQ-5D-5L for women and residents with longer length of stays. There was no significant between-care home influence on either ASCOT-Proxy-Resident or ICECAP-O. For EQ-5D-5L, there were between-care home level associations, with resident functional dependence, moderate and moderate severe cognitive impairment and for residents aged 80-89 years having a positive influence on health-related QoL between care homes.

### Construct validity by hypothesis testing

Evaluation of the hypotheses concerning differences in QoL scores based on personal and care home characteristics are presented in Table 4. There was sufficient evidence of construct validity for all three QoL measures as >=75% of hypotheses were accepted.

**Table 4:**
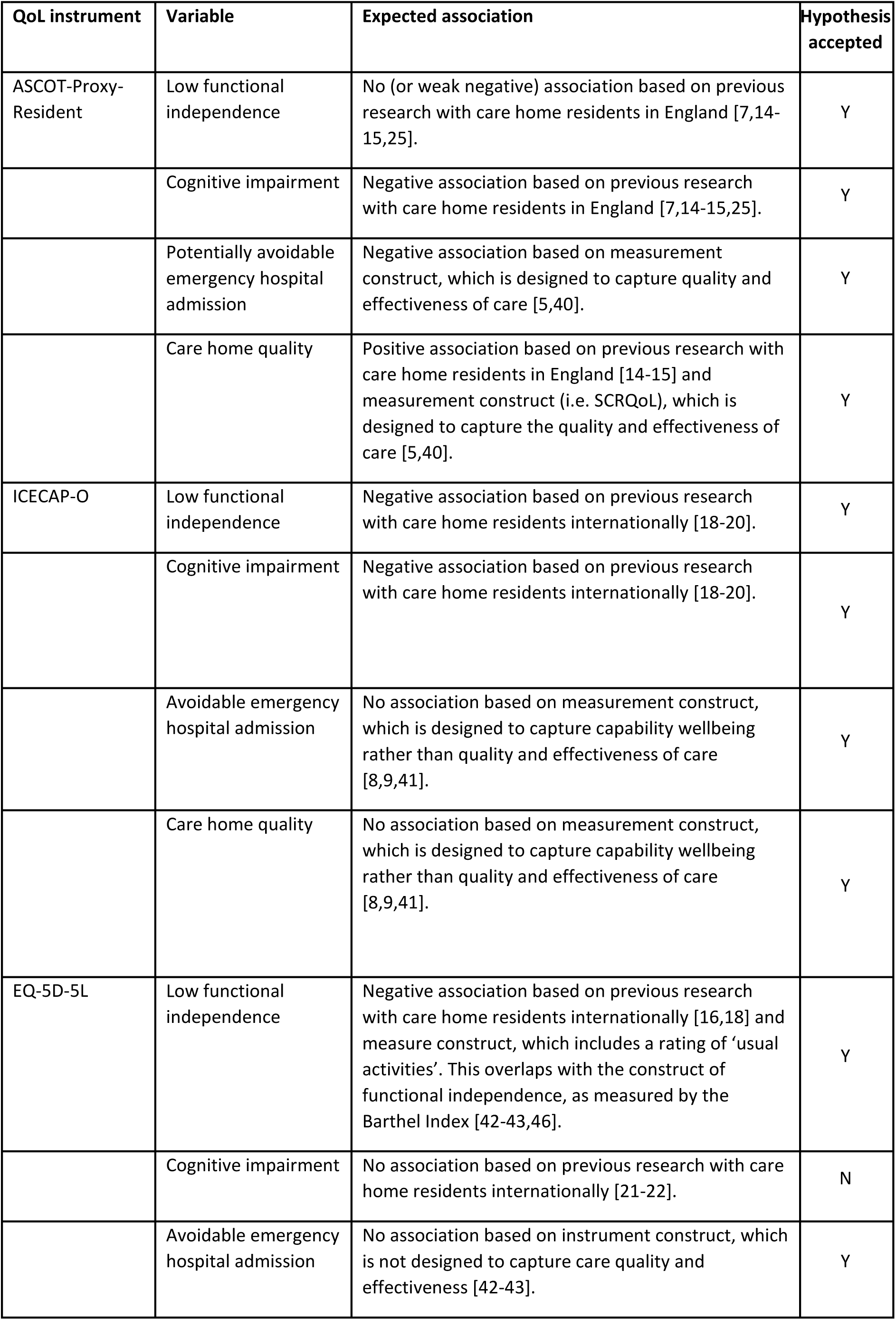
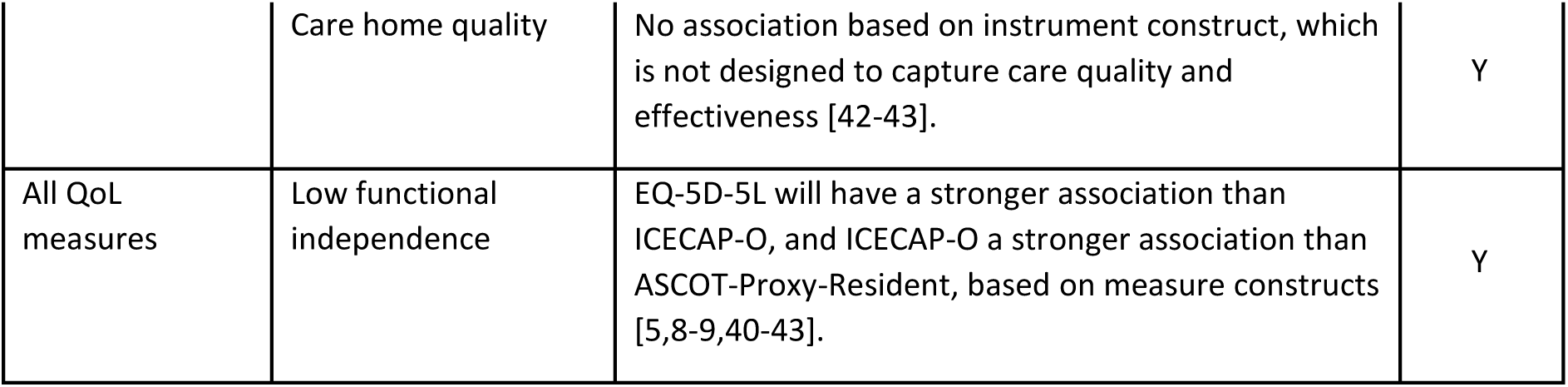
Construct validity hypotheses, by QoL measure.

## Discussion

This study looked to assess the construct validity of three different QoL measures, namely the ASCOT-Proxy-Resident, ICECAP-O and EQ-5D-5L Proxy 2, using discriminant analysis and to understand factors associated with the QoL of care home residents in England. The findings showed that each measure is associated with different factors contributing to QoL. EQ-5D-5L was most strongly associated with dependence in performing activities of daily living and also with the highest level of cognitive impairment but not with avoidable hospital admissions or comorbidities. A lack of association with cognitive function in the older people has been previously established [21-22] and, whilst EQ-5D-5L is well established as a measure of health-related QoL, it also does not appear to differentiate between care home residents regarding comorbidity.

As such, for care home studies, particularly those looking to establish the impact of interventions on overall wellbeing, it would be beneficial to measure QoL more broadly using the ASCOT-Proxy-Resident and ICECAP-O. ICECAP-O can differentiate between residents by level of functional dependence; both measures were able to differentiate residents by level of impairment and also between residents with and without delirium. Further, ASCOT-Proxy-Resident can differentiate by level of care quality and effectiveness indicators, whilst controlling for functional dependence. This is both at the care home level, as in previous analyses using ASCOT CH4 [14,15], and also at resident level with a negative association with avoidable emergency hospital admissions. Hospital admissions are likely for care home residents [27] and decisions behind them are complicated, being affected by several factors [60]. However, we expect that, other things equal, an admission classed as potentially avoidable would be more likely to be indicative of some form of unmet need for a care home resident, be that health- or social care-related. The modelling strategy used in this study also confirms that the finding is not an indicator of care home-level quality, but rather relates to individual-level care in relation to the resident’s fluctuating needs. However, more work is needed to analyse the impact that appropriate health care utilisation has on care home resident QoL using a longitudinal approach.

Overall, this study has added to the literature by analysing three instruments of QoL and their association with personal and health characteristics and care home level factors. The study has shown that all three QoL measures have acceptable construct validity for use in care homes, which adds to other evidence from the DACHA study on the psychometric properties of the measures [26,40], and demonstrates that each measure captures different constructs, indicating that the measures are complementary rather than duplicative. For ASCOT-Proxy-Resident, the DACHA study is the first time that this measure have been used within care homes in England. The study has also added to the literature by assessing the association between resident QoL and avoidable emergency hospital admissions. Further, the findings are in line with previous recommendations that economic evaluations of older people, including those living in care homes, should use different, complementary, QoL measures to consider different constructs, e.g. health-related and social care-related QoL [12-13]. Our findings here support the inclusion of multiple measures of different constructs (health-related QoL, social care-related QoL, capability wellbeing) in a future MDS, subject to the additional data burden for care home providers and their staff [61].

The study has demonstrated the value of the DACHA MDS pilot data, especially in the collection of resident QoL. Likewise, any future MDS must have consistent data and be of value to residents [61]. As such, data in a future MDS would enable research to inform national, local and provider policy and care delivery to improve resident QoL. For example, with relevant data, research could analyse the impact of health care utilisation [16] and staffing [62-63] on resident QoL. It could also be used to assess the effectiveness of care, for example, comparing models of care, and provide evidence on the value for money of residential care [64]. Importantly, data in any future MDS would enable analyses to appropriately control for the health and social care needs of residents.

There are a number of limitations to this study. First, the findings should not be seen as representative of the English care home resident population, although for ASCOT-Proxy-Resident, the profile of social care-related QoL is in line with past research that used a mixed-methods approach to collect social care-related QoL [14-15], and they are also similar to findings for ICECAP-O and EQ-5D-5L internationally [17-22]. Second, we were unable to assess changes to QoL over time due to issues with data quality in a second wave of data collection for the pilot MDS [35]. The level of needs of a care home resident are likely to increase over time [65], and it would be of interest to assess how QoL changes with this. Currently, longitudinal analysis is limited in England to those living in their own homes [66-67] or a specific group of residents [68]. A longitudinal analysis would also help mitigate concerns of bias due to omitted variables, which, given the statistical methods employed, was still possible at the resident level [58]. Finally, there were no homes with an overall CQC rating rated as ‘Inadequate’ included in the study. However, the ‘Inadequate’ rating is transitory in nature, with care homes having to improve or face closure [69].

## Conclusion

This study has found that the ASCOT-Proxy-Resident, ICECAP-O and EQ-5D-5L Proxy 2 have acceptable construct validity for use in care homes. Findings also evidence that they are complementary measures based on different constructs. In so doing, the study has demonstrated both the value of the DACHA study pilot MDS data, especially in collecting resident QoL, and a rationale for the inclusion of these three QoL measures in any future MDS.

## Data Availability

Anonymised data extracted from digital care records will be available on request from the corresponding author, SA, following a 24 month embargo from the date of publication.

## Funding statement

This project is funded by the National Institute for Health Research (NIHR) Health Service Research and Delivery programme (HS&DR NIHR127234) and supported by the NIHR Applied Research Collaboration (ARC) East of England.

AMT, AG, KS, AK and CG are supported by the NIHR Applied Research Collaborations in Kent, Surrey and Sussex; East Midlands; Yorkshire and Humber and East of England, respectively.

AG, KS and CG are NIHR Senior Investigators.

The views expressed are those of the authors and not necessarily those of the NIHR or the Department of Health and Social Care.

## Acknowledgments

We would like to acknowledge and thank the Public Involvement and Engagement Panel members, software providers, care homes, care staff and residents for participating in the study, as well as the wider DACHA study team whose work informed the development of the DACHA (Developing resources And minimum dataset for Care Homes’ Adoption) Minimum Data Set and contributed to the selection of the quality of life measures included in the pilot.

